# Rapid Screening of COVID-19 Disease Directly from Clinical Nasopharyngeal Swabs using the MasSpec Pen Technology

**DOI:** 10.1101/2021.05.14.21257006

**Authors:** Kyana Y. Garza, Alex Ap. Rosini Silva, Jonas R. Rosa, Michael F. Keating, Sydney C. Povilaitis, Meredith Spradlin, Pedro H. Godoy Sanches, Alexandre Varão Moura, Junier Marrero Gutierrez, John Q. Lin, Jialing Zhang, Rachel J. DeHoog, Alena Bensussan, Sunil Badal, Danilo Cardoso, Pedro Henrique Dias Garcia, Lisamara Dias de Oliveira Negrini, Marcia Ap. Antonio, Thiago C. Canevari, Marcos N. Eberlin, Robert Tibshirani, Livia S. Eberlin, Andreia M. Porcari

## Abstract

The outbreak of COVID-19 has created an unprecedent global crisis. While PCR is the gold standard method for detecting active SARS-CoV-2 infection, alternative high-throughput diagnostic tests are of significant value to meet universal testing demands. Here, we describe a new design of the MasSpec Pen technology integrated to electrospray ionization (ESI) for direct analysis of clinical swabs and investigate its use for COVID-19 screening. The redesigned MasSpec Pen system incorporates a disposable sampling device refined for uniform and efficient analysis of swab tips via liquid extraction directly coupled to a ESI source. Using this system, we analyzed nasopharyngeal swabs from 244 individuals including symptomatic COVID-19 positive, symptomatic negative, and asymptomatic negative individuals, enabling rapid detection of rich lipid profiles. Two statistical classifiers were generated based on the lipid information aquired. Classifier 1 was built to distinguish symptomatic PCR-positive from asymptomatic PCR-negative individuals, yielding cross-validation accuracy of 83.5%, sensitivity of 76.6%, and specificity of 86.6%, and validation set accuracy of 89.6%, sensitivity of 100%, and specificity of 85.3%. Classifier 2 was built to distinguish symptomatic PCR-positive patients from negative individuals including symptomatic PCR-negative patients with moderate to severe symptoms and asymptomatic individuals, yielding a cross-validation accuracy of 78.4% accuracy, specificity of 77.21%, and sensitivity of 81.8%. Collectively, this study suggests that the lipid profiles detected directly from nasopharyngeal swabs using MasSpec Pen-ESI MS allows fast (under a minute) screening of COVID-19 disease using minimal operating steps and no specialized reagents, thus representing a promising alternative high-throughput method for screening of COVID-19.

## Introduction

The novel coronavirus disease 2019 (COVID-19), caused by the severe acute respiratory syndrome coronavirus 2 (SARS-CoV-2), has presented an unprecedented global challenge to society and public health^1,2^. As vaccines have yet to be widely administered to the public, specially in resource-limited countries, and their effectiveness towards new variants are yet to be determined, mitigation of disease transmission relies heavily on the widespread availability of rapid COVID-19 tests exhibiting robust analytical performance and diagnostic metrics including adequate sensitivity, specificity, and low false-positive rates (FPR) and false-negative rates (FNR)^3,4^. Current diagnostic assays for COVID-19 are largely based on the detection of SARS-CoV-2 ribonucleic acid (RNA) via quantitative polymerase chain reaction (qPCR) analysis. PCR is a powerful and highly sensitive assay; yet, clinical laboratories have faced challenges in maintaining current demands due to limited availability of the specialized test reagents, instrumentation that have been overrun beyond their capabilities, and low throughput analyses^5^. Alternative diagnostic tests that have little to no reagent requirements and provide a rapid turnaround time are highly valuable for COVID-19 detection^6^. For example, serological tests targeting host antibodies have been deployed for COVID-19 diagnosis, yielding promising results^7,8^. Yet, the inability to diagnose early-stage or acute infections with antibody testing, along with potential cross-reactivity from prior infections by other pathogens presents a challenge for patient screening^9^. Antigen tests have been developed to rapidly identify active SARS-CoV-2 infections via the detection of the nucleocapsid protein antigen^6^. While antigen tests provide diagnosis in ∼15 min, FNR of up to 40% have been reported due to higher limits of detection compared to PCR^10-12^. Alternative testing and screening methods capable of rapidly screening for COVID-19 disease are thus still needed to increase testing capacity and throughput.

COVID-19 tests targeting molecular species other than viral RNA are currently being evaluated as rapid screening methods prior to PCR analysis to mitigate viral outbreaks. Lipids present an interesting molecular target for identifying SARS-CoV-2 infection as these molecules are a major component of the viral envelope and are involved in key replication cycle processes, including the production of new virions^13,14^. Viral genetic material does not code for lipids but sequesters these molecules from their host cellular membranes during budding. The lipid composition of the host-derived viral envelope is known to be specific to the budding site^15^ and quantitatively distinct from that of the host membrane and from other viruses^16-19^. Coronaviruses, for example, bud and derive their viral envelope lipids from the membrane of the host endoplasmic reticulum (ER)-Golgi intermediate complex^16^, whereas the influenza virus acquire their lipids from the host apical plasma membrane^20^. In a study by Van Genderen *et al*, the proportion of phosphatidylinositol (PI) in the viral membranes of coronavirus murine hepatitis virus (MHV) was found to be elevated by 4% compared to the host cells, and the ratio of phosphatidylserine (PS) to PI species was reduced by 12%^21^. Viral pathogens also remodel host lipid metabolism to enable replication during infection, altering the overall lipid composition of infected host cells. For example, Yan *et al* described that fatty acids and glycerophospholipids were significantly elevated in human cells infected with the H-CoV 229E coronavirus compared to healthy cells^22^. Dysregulation of highly abundant glycerophospholipids in infected host cells and the unique lipid composition of the pathogen itself, therefore, represent a promising target for diagnostic tests.

Mass spectrometry (MS) technniques have been largely applied to study infectious diseases, targeting various biological molecules to identify bacterial and viral infections^23-27^. Recently, MS techniques have been explored to detect COVID-19 based on metabolite, lipid, and protein information^28-34^. For example, liquid chromatography MS and machine learning models have been used to identify proteomic and metabolic signatures in sera from COVID-19 patients with 93.5% accuracy for a training set of 31 samples^31^. Matrix-assisted laser desorption/ionization mass spectrometry (MALDI-MS) was also used to analyze extracts of nasal swabs^29^ and plasma^35^ to diagnose COVID-19. Ambient ionization MS and machine learning have been explored to detect SARS-CoV-2 infection based on fatty acid and lipid profiles^30^. De Silva *et al* used paper spray MS to analyze lysed cell extracts from 30 symptomatic COVID-19 positive and symptomatic negative patients, with 93.3% agreement with PCR based on 11 metabolites, fatty acids, and lipids^30^ whereas Ford *et al* used desorption electrospray ionization (DESI) and laser desorption rapid evaporative ionization mass spectrometry (LD-REIMS) to analyze 70 nasal swabs, with accuracies over 84.0%^32^. As MS technologies steadily advance towards clinical implementation, these studies showcase the potential of MS-based assays for screening and diagnosis of viral infections.

Here, we report a new design of the MasSpec Pen technology for the analysis of swabs and demonstrate its use for rapid and direct lipid analysis and potential for COVID-19 screening. We previously reported the development of the MasSpec Pen as a handheld device integrated to a mass spectrometer for direct and rapid molecular analysis of tissues^36,37^. While the handheld MasSpec Pen was designed as an easy-to-use device that enabled precise and efficient molecular analyses of sample surfaces using a solvent droplet, it precludes sufficient sampling and full area coverage of three-dimensional samples such as swab tips that contain heterogeneous adhesion and distribution of mucous secretion. To that end, we optimized the disposable device to enable uniform sampling of an entire swab tip via liquid extraction using common solvents, and thus efficient molecular extraction and analysis. The disposable sampling system was then directly integrated to an ESI source for sensitive detection of molecular ions. Using the MasSpec Pen-ESI MS system, we obtained rich lipid profiles from nasopharyngeal swabs and built statistical classification models to evalulate its predicition capabilities for COVID-19. Collectively, our study shows that direct analysis of clinical swabs with the MasSpec Pen-ESI MS technology is a potentially promising method for rapid screening of viral infections such as COVID-19.

## Materials and Methods

### Chemicals

Cardiolipin (CL) 72:4, phosphatidylglycerol (PG) 36:2, and phosphatidylethanolamine (PE) 36:2 lipid standards were purchased from Avanti Polar Lipids (Alabaster, AL). PG and PE standards were dissolved in methanol at a concentration of 10 µM, and the CL standard was dissolved in methanol at a concentration of 13 µM.

### Design and Fabrication of the adapted MasSpec Pen-ESI system

The MasSpec Pen polydimethylsiloxane (PDMS) swab sampling device was designed in CAD software and negative molds for the devices were fabricated using procedures previously described (Methods and Supplementary Materials)^37^. The sub-atmospheric pressure ESI source was built by modifying the housing of a commercially available atmospheric pressure chemical ionization (APCI) source (Agilent Technologies). See **Supplementary Information** for details on the modifications made to the APCI source and materials used to make the lab-built sprayer.

### Clinical Nasopharyngeal Swabs

Nasopharyngeal swabs from symptomatic SARS-CoV-2 PCR positive and symptomatic and asymptomatic SARS-CoV-2 PCR negative individuals were collected from consented patients that were hospitalized with moderate or severe respiratory symptoms in two different hospitals (Santa Casa and Bragantino) as well as from asymptomatic volunteers at Integrated Unit of Pharmacology and Gastroenterology (UNIFAG) in the city of Bragança Paulista (São Paulo, Brazil), by the research team at the University of San Francisco (Bragança Paulista, Sao Paulo, Brazil). Approval from Institutional Review Board (IRB) was received for the study (protocol number 31573020.9.0000.5514, approved from May 29, 2020). Additional details about clinical swab collection, handling, and storage are in the **Supplementary Information**.

Clinical sample collection began on July 17, 2020. Clinical diagnosis for the symptomatic patients and asymptomatic individuals was performed via RT-PCR analysis using a different clinical swab as part of their clinical care and independently of our study. **Table 1** provides patient demographics information. As of October 21, 2020, swabs from 268 individuals have been collected in Brazil and shipped to and received by our laboratory at UT Austin where swabs were stored at -80°C prior to analysis.

**Table 1.** Patient demographic information for the samples used in Lasso statistical analysis. Data are presented as medians (interquartile ranges, IQR) or quantity of categorical variables (%). The demographic information is for all patients and individuals included in the Lasso statistical analysis. The clinical information is for the symptomatic negative and symptomatic positive patients included in the statistical analysis.

| Parameters | Symptomatic<br>PCR Positive | Symptomatic<br>PCR Negative | Asymptomatic<br>PCR Negative | p-value |
| --- | --- | --- | --- | --- |
| **Demographics** |  |  |  |  |
| Number of patients, n | 44 | 26 | 101 | 2.18E-06 |
| Age range, y | (21, 84) | (26, 84) | (20, 89) |  |
| Sex (female, male) | (16, 28) | (11, 15) | (58, 43) |  |
| **Symptoms** |  |  |  |  |
| Fever | 19 | 12 | - | 1.0000 |
| Cough | 30 | 16 | - | 0.7602 |
| Myalgia | 9 | 2 | - | 0.2811 |
| Sore throat | 8 | 8 | - | 0.3590 |
| Headache | 12 | 1 | - | 0.0342 |
| Dyspnea | 29 | 18 | - | 0.9820 |
| Tiredness/fatigue | 3 | 2 | - | 1.0000 |
| Loss of smell/taste | 9 | 9 | - | 0.3045 |
| Diarrhea | 11 | 2 | - | 0.1386 |
| None | - | - | 101 | - |
| **Chest CT Features** |  |  |  |  |
| Ground-glass opacity | 42 | 18 | - | 0.0024 |
| Consolidations | 21 | 14 | - | 0.8770 |
| Crazy paving appearance | 21 | 9 | - | 0.3659 |
| Reticular pattern | 6 | 6 | - | 0.5214 |
| Pulmonary commitment degree | 37 | 16 | - | 0.0410 |
| Suggestive of viral infection | 42 | 18 | - | 0.0024 |
| **Underlying conditions** |  |  |  |  |
| Systemic arterial hypertension | 25 | 11 | 16 | 0.5765 |
| Cardiovascular disease | 6 | 4 | 2 | 1.0000 |
| Obesity | 10 | 1 | 22 | 0.0788 |
| Diabetes | 16 | 2 | 3 | 0.0178 |
| Lung disease | 4 | 5 | 8 | 0.3925 |
| Chronic obstructive pulmonary diesase | 1 | 1 | 1 | 1.0000 |
| Smoker/ex-smoker | 3 | 3 | 6 | 0.8105 |
| Asthma | 2 | 2 | 2 | 0.9879 |

### MasSpec Pen-ESI Analysis and Data Acquisition

Prior to the analysis, the swabs were removed from the -80°C freezer and thawed to room temperature in a Class II biological safety cabinet for 15 min. To maximize safety measures, swabs were then heat-inactivated for 30 min at 65°C. Following heat inactivation, swabs were placed in the biological safety cabinet until cooled to room temperature. Swabs were stored in a refrigerator at 4°C until MasSpec Pen-ESI MS analysis. Swabs were analyzed within 3 days of heat inactivation.

Experiments were performed on two mass spectrometers, an LTQ Orbitrap XL and a Q Exactive HF mass spectrometer (Thermo Fisher Scientific). The **Supplementary Information** provides experimental details and parameters. During MasSpec Pen-ESI MS analysis, the swab tips were inserted into the middle channel of the PDMS sampling device (**Figure 1**). Upon the press of a foot pedal, a volume of 167 µL of CHCl_3_:MeOH is delivered from the syringe pump to the middle channel containing the swab, interacting with and extracting molecules from the swab tip for 10 s. The entire process is controlled using programmed microcontrollers. A vacuum was then applied for 30 s to the PTFE tube to enable the transport of the solvent from the swab reservoir to the ESI source. Mass spectra were acquired for ∼20-30 s.

**Figure 1.**
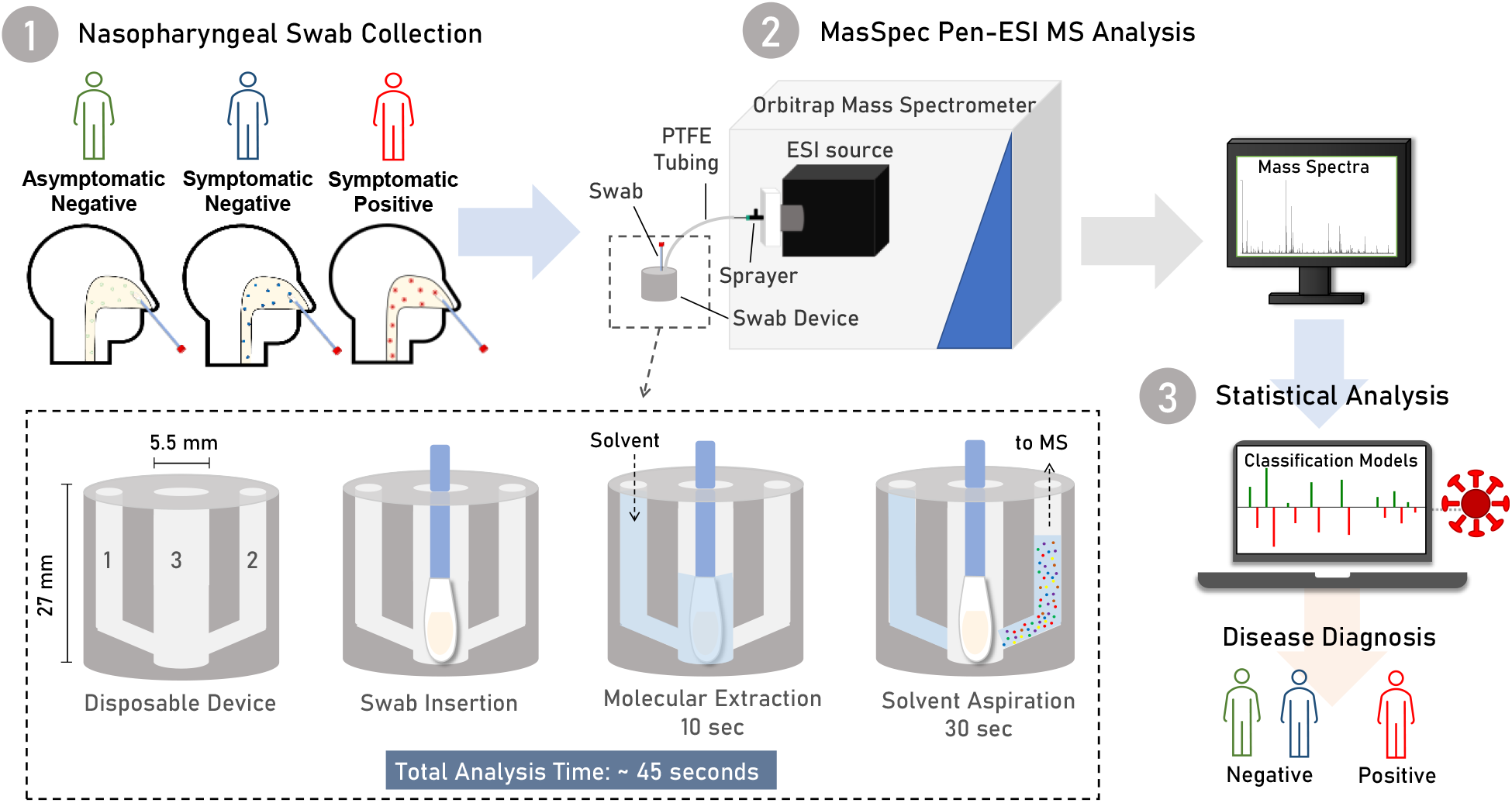
Schematic of the MasSpec Pen-ESI-MS system for the diagnosis of COVID-19 infection. Swabs acquired from symptomatic patients and asymptomatic individuals were analyzed by the MasSpec Pen-ESI MS platform and the mass spectra collected were used to build machine learning classification models for diagnosis of COVID-19. A zoom view of the design MS swab device and the steps for analysis. The insert shows two conduits for incoming solvent (1) and aspiration of the solvent containing the extracted molecules (2) and a middle reservoir (2). During analysis, the swab is inserted into the middle reservoir. Upon the press of a foot pedal, solvent is delivered to the middle reservoir to interact with the swab tip to extract molecules. After 10 sec the solvent containing the extracted molecules is transported the mass spectrometer for ESI analysis.

### Statistical Analysis

Seventy-five mass spectra were averaged and extracted for each sample analyzed. A mass filter of *m/z* 400-1000 was applied, after binning and background subtraction. Data was normalized to the TIC, and peaks appearing in less than 50% of the entire data set for each classifier were removed during cross validation (CV). The least absolute shrinkage and selection operator (Lasso) statistical analysis was performed using the beta version of the glmnet package v4.1-2, using the exclude/filtering option in glmnet. The time elapsed between PCR and MS swab collection and days since symptom onset were used as selection criteria. Swabs that were collected for MS three days or more after PCR sample collection and beyond 14 days of symptom onset were excluded from the classifier. From the 268 individuals who had their swab samples collected, 171 met the criteria of the time interval to PCR-sample collection and days from symptoms onset. Demographic and clinical information for the patients from which the selected samples were obtained is provided in **Table 1 and S1**. Based on the selection criteria, two statistical classifiers were generated: 1) asymptomatic COVID-19 PCR negative (n= 101) vs COVID-19 symptomatic PCR positive (n=44) samples and 2) COVID-19 PCR negative (n=101 asymptomatic and n=26 symptomatic) vs COVID-19 symptomatic

PCR positive. 10-fold CV was used with Lasso to generate predictive models. For Classifier 1, the data was randomly split into a training set (2/3 of data, n=97) and a validation set (1/3 of data, n=48). Additionally, we tested Classifier 1 on a withheld test set of PCR-negative symptomatic samples (n=26). The performance of the models were evaluated by measuring the predictive accuracy, sensitivity, specificity, negative predictive value (NPV), and positive predictive value (PPV) which were calculated based on the agreement with PCR diagnosis. Statistical analysis were performed by K.Y.G., M.F.K., J.Q.L. and J.M.G., and independently verified by R.T.. The **Supplementary Information** describes detailed data preprocessing, inclusion criteria for statistical analysis, and details on Lasso and other statistical methods.

## Results

### Design of the MasSpec Pen-ESI System

We previously developed the MasSpec Pen as a handheld device directly coupled to a mass spectrometer for direct analysis of tissues^37^. The PDMS pen tip was comprised of a solvent reservoir that held a solvent droplet in contact with a sample surface to enable efficient molecular extraction. While this design is intuitive for handheld use and well-suited for the analysis of tissue regions, the area covered by the reservoir opening (typically ∼5.7 mm^2^) was insufficient for uniform sampling and analysis of the secretion covering the three-dimensional area of an entire swab tip. To enable sensitive and robust analysis of all the mucous secretion material in and on a swab tip, we thus redesigned and optimized the MasSpec Pen device, interface, and ionization system with the goal of ensuring direct and efficient sampling of the entire swab tip while maintaining the ease of use and rapid nature of the analysis of the original MasSpec Pen. The PDMS swab sampling device was integrated via PTFE tubing with a sub-atmospheric pressure ESI source for effective ionization and sensitive analysis of the extracted molecules (**Figure 1)**. Similar to the original MasSpec Pen PDMS tip, the PDMS sampling unit is designed with three conduits that connect to a middle reservoir that was widened to 5.5 mm diameter and ∼22 mm height to enable an entire swab tip to be fully inserted (**Figure 1 insert**). A PTFE tube connected to a syringe pump was then inserted into conduit 1 for the delivery of solvent to the middle reservoir, whereas a second PTFE tube was inserted into conduit 2 for solvent aspiration into the sub-atmospheric pressure ESI source. The swab analysis is then performed with minimal operational steps: after the swab is inserted into the middle swab reservoir, solvent is delivered to the reservoir through a PTFE tube connected to conduit 1 via the press of a foot pedal, where the solvent interacts with the entire swab tip for 10 s for analyte extraction. Following the extraction period, conduit 2 is opened for 30 s allowing the vacuum to aspirate solvent to the sprayer for ESI analysis. Within the ESI source, a lab-built sprayer promotes ionization of the molecules extracted within the solvent. Note that a sub-atmospheric pressure was set within the ESI housing, measured via the forevacuum pressure of the mass spectrometer (1.4-1.6 mbar), with the sole purpose of enabling suction and thus transport of the solvent from the swab reservoir to the ESI sprayer.

### Optimization of the MasSpec Pen-ESI system for swab analysis

Using the MasSpec Pen-ESI system coupled to an LTQ XL mass spectrometer, we first evaluated commonly used medical polyester and nylon flock swabs sterilized with ethylene oxide or gamma irradiation for assay compatibility by dipping the swabs in a solution of cardiolipin (CL) 72:4 standard followed by analysis using MasSpec Pen-ESI MS. For the analysis of nylon flock swabs sterilized using ethylene oxide, a series of interfering ions identified as repeating units of ethylene oxide were observed from *m/z* 350-1200 at a ∼5.5 fold (n=5, *m/z* 735.420) higher relative abundance compared to the CL standard ion of *m/z* 727.510 (**Figure S1**), thus hindering the detection of the CL standard due to ion suppression. Yet, no polymer ions were observed in the mass spectra obtained from polyester flock swabs sterilized with gamma irradiation. Thus, all consecutive experiments including collection and analysis of clinical samples were therefore performed with polyester flock swabs sterilized by gamma irradiation to avoid polymer interference.

Next, solvent composition and the volume used to fill the reservoir were optimized to ensure the entire swab was saturated with solvent to efficiently extract molecules, as well as allow for consistent signal and ESI spray stability during the analysis. Different organic solvent systems with volumes ranging from 100 µL to 200 µL were evaluated, with a solvent volume of 167 µL selected as optimal. Among the solvent systems tested, CHCl_3_:MeOH (1:1, v/v) yielded the highest reproducibility (relative standard deviation of 6.4% (n=10)) and spray stability (∼20-30 s of ion signal) while minimizing the extraction and detection of interfering ions. Notably, a 25.5% (n=4) increase in signal intensity of lipids was achieved when compared to the traditional MasSpec Pen setup^38^ (**Figure S2**), likely due to more efficient ionization and desolvation provided by ESI. Altogether, a total analysis time of 45 s or less per swab was achieved, which included 5 s of solvent delivery, 10 s of swab extraction time, and ∼20-30 s droplet transport and ESI signal (**Figure 1**). Lastly, we also evaluated if heat inactivation led to any substantial change or degradation to the lipids contained in the sample using PG and PE lipid standards (**Figure S3**). We found no statistical significance (p>0.05) in the mean intensity of the lipids detected from the heat-inactivated or control swabs, indicating that the inactivation process did not significantly alter lipid composition.

### Molecular analysis of clinical nasopharyngeal swabs

As **Figure 2a** shows, we observed rich molecular profiles composed of a diverse range of glycerophospholipid and lysolipid species in mass spectral profiles of symptomatic COVID-19 positive and negative swabs and asymptomatic healthy samples. Note that the *m/z* 400-1000 range was used to avoid detection and ion suppression from non-biological interferents detected as ions of *m/z* <400 while enabling detection of a broad range of lipid species. Various ions tentatively identified lysoPE 16:0 (*m/z* 452.278), lysoPE 18:0_0:0 (*m/z* 480.309), lysoPC 18:1 (*m/z* 556.318) (**Figure 2B**), and cholesterol sulfate (*m/z* 465.304) were observed in the *m/z* range 400-600. Additionally, molecules such as ceramide species including Cer 34:1 (*m/z* 572.481), Cer 36:1 (*m/z* 601.533), and Cer 42:2 (*m/z* 682.591) as well as glycerophospholipids including PS 18:1_18:0 (*m/z* 788.545), PI 20:4_18:0 (*m/z* 885.550), and PE 34:2 (*m/z* 714.509) were observed (**Figure 2C**).

**Figure 2.**
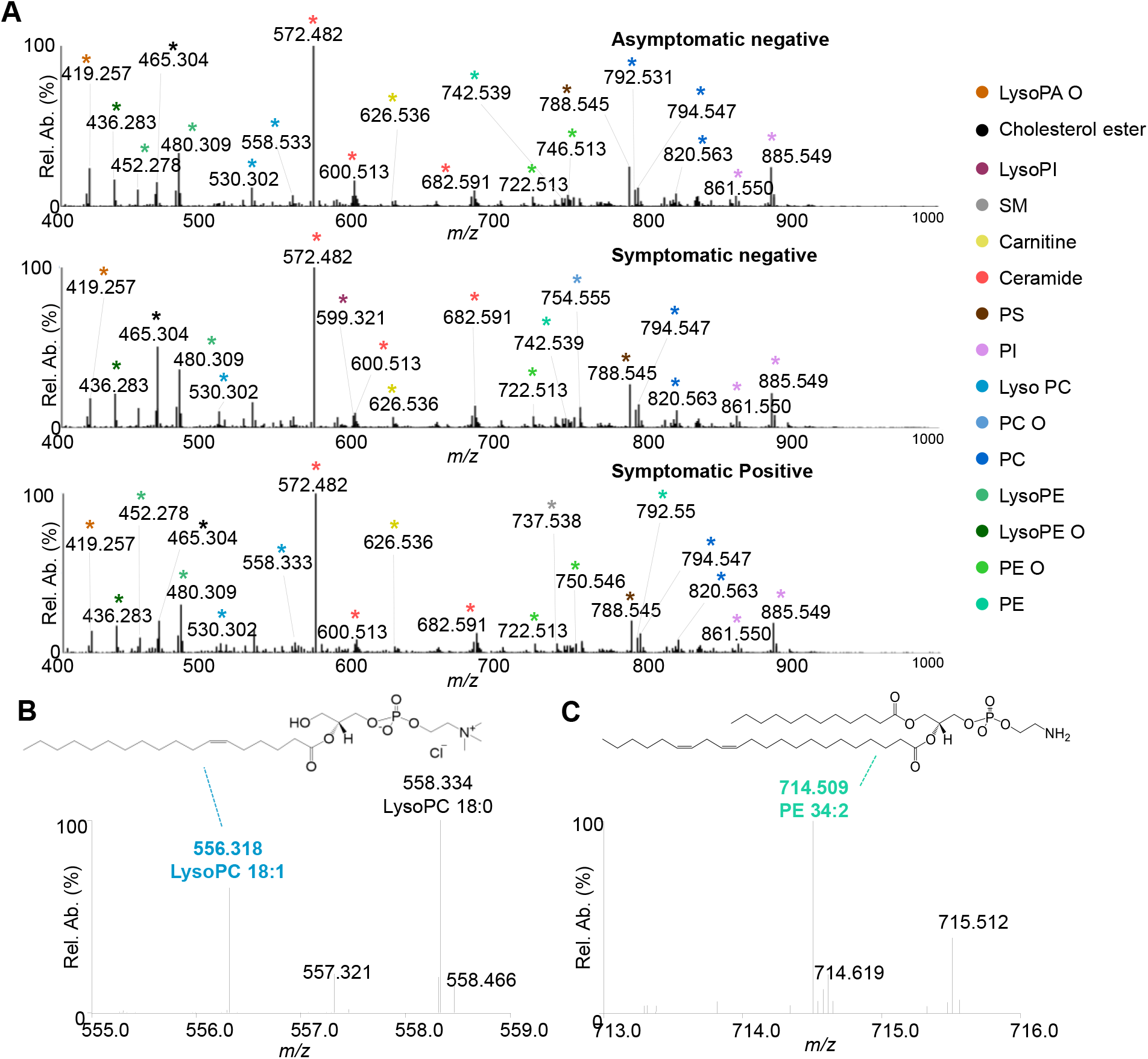
MasSpec Pen-ESI MS analysis of asymptomatic negative, symptomatic negative, and symptomatic positive samples. **A)** Averaged spectra of all asymptomatic negative (n=101, top), symptomatic negative (n=26, middle), and symptomatic positive (n=44, bottom). Different colored peaks correspond to different lipid classes which are labeled in the legend. **B)** Zoom in of *m/z* range 555-559 to show the detection of lysoPC species including lysoPC 18:1 (*m/z* 556.318) and 18:0 (*m/z* 558.334) with the structure for lysoPC 18:1 shown in a mass spectra from a positive sample. **C)** Zoom in of *m/z* range 713-716 to show the detection of PE 34:2 (*m/z* 714.509) and the corresponding structure in a mass spectra from a positive sample.

### Statistical prediction of COVID-19 infection

We next statistically evaluated if the molecular information obtained with the MasSpec Pen-ESI MS system was predictive of COVID-19 infection. We first employed the Lasso method to build a classification model to discriminate data obtained from symptomatic patients positive for COVID-19 (n=44) from asymptomatic individuals negative for COVID-19 (n=101), termed **Classifier 1**. The model exhibited a strong performance using 10-fold CV (n=97), yielding an area under the receiver operating characteristic (ROC) curve (AUC) of 0.852 (**Figure 3A**) and an accuracy of 83.5% (**Figure 3C**). A prediction probability value of 0.350 was selected as optimal threshold value for sample classification based on the ROC curve. Samples with a probability lower than 0.350 were classified as asymptomatic negative and those with a probability higher than 0.350 were classified as symptomatic positive (**Figure 3B**). Using this approach, a total of 58 out of 67 asymptomtic negative and 23 out of 30 symptomatic positive samples had a prediction result in agreement with PCR, resulting in 76.7% sensitivity and 86.6% specificity (**Figure 3C**). We also calculated the NPV and PPV to evaluate the ability of our statistical classifier to provide a predictive COVID-19 diagnosis that aligns with the true absence or presence of the disease. The model yielded an NPV of 89.2% and a PPV of 71.9%.

**Figure 3.**
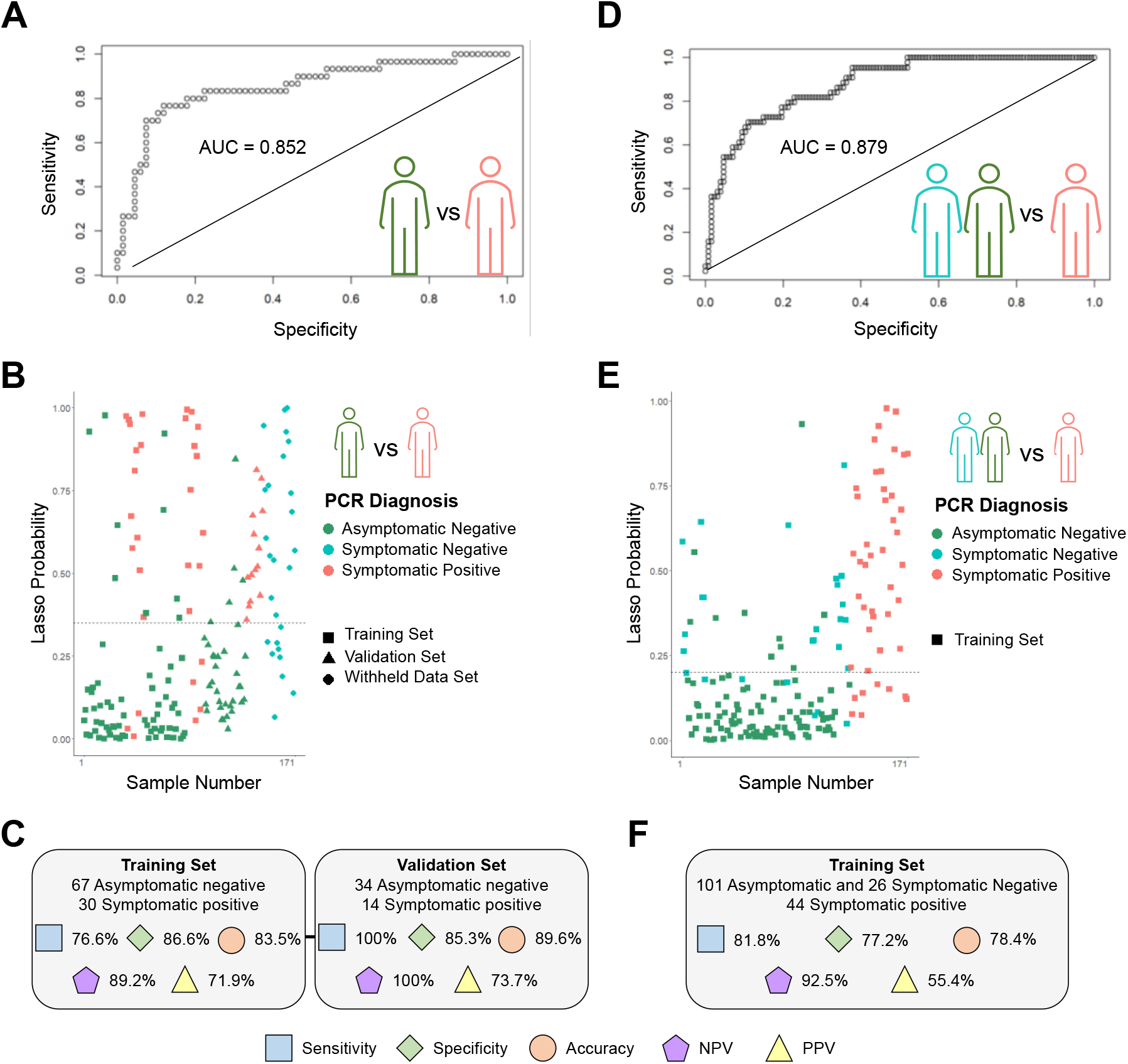
Statistical analysis results for Classification models 1 and 2. Classifier 1, asymptomatic negative vs symptomatic positive, **A)** ROC curve, **B)** Plot of the classification probabilities for samples used in the training and validation set. The dashed line represents the cutoff value for classification as asymptomatic negative or symptomatic positive for COVID-19 (0.350). **C)** Sensitivity, specificity, accuracy, NPV, and PPV and for the training and validation set for Lasso Classifier 1. Classifier 2, negative vs positive, **D)** ROC curve, **E)** Plot of the classification probabilities for samples used in the training for Classifier 2. The dashed line represents the cutoff value for classification as negative or positive for COVID-19 (0.201). **F)** Sensitivity, specificity, accuracy, NPV, and PPV for the training set for Lasso Classifier 2.

Next, we assessed the predictive performance of **Classifier 1** using a validation set of 34 asymptomatic negative and 14 symptomatic positive samples. Only five asymptomatic negative samples were classified as positive in disagreement with PCR while all symptomatic positive samples were correctly classified, resulting in an overall agreement with PCR of 89.6%, a specificity of 85.3%, sensitivity of 100%, NPV of 100%, and PPV of 73.7%. We then used the classifier to predict on a withheld set of samples obtained from patients presenting respiratory symptoms similar to those associated with COVID-19 disease (n=26) but that had a negative PCR result (**Table S1**). A total of nine samples in the withheld set of data were classified as negative in agreement with PCR, whereas 17 samples were classified as positive, in disagreement with the PCR diagnosis. Out of these 17 patients, 12 had a chest computational tomography (CT) that was suggestive of viral infection, presenting ground-glass opacity (GGO) among other features such as consolidation and pulmonary commitment^39^. **Table S2** provides a detailed summary of the classification results.

To more broadly evaluate performance for COVID-19 screening, we built a second classifier, termed **Classifier 2**, that combined the samples from the symptomatic negative patients (n=26) and the asymptomatic negative individuals (n=101) into a single negative class, whereas the positive class was comprised of samples from symptomatic positive patients (n=44). The predictive model was comprised of 41 *m/z* features and yielded an overall agreement with PCR of 78.4%, sensitivity of 81.8%, specificity of 77.2% (**Figure 3F**), and an AUC of 0.879 (**Figure 3D**). A prediction probability threshold of 0.201 was selected to maximize the sensitivity of **Classifier 2** (**Figure 3E**). A total of 29 PCR negative samples were classified as positive by our method, among which 19 were from symptomatic patients that presented respiratory symptoms similar to those associated with COVID-19 disease. We also noted that 13 of the 19 symptomatic negative samples classified as positive had chest CT results suggestive of viral infection.

Among the selected features used to build the several classification models, various lipids were selected to discriminate COVID-19 positive disease and negative diagnosis. For example, several PE and lysoPE species were selected as important for predicting negative status among the two classifiers generated, including lysoPE 20:0 (*m/z* 508.341), and PE 34:2 (*m/z* 714.508), (**Figure 4**). For **Classifier 1**, various ceramides such as Cer 42:3 (*m/z* 680.576), Cer 42:2 (*m/z* 682.591), Cer 43:3 (*m/z* 694.592), Cer 44:5 (*m/z* 700.587) were selected as characteristic of symptomatic positive COVID-19 infection by Lasso, whereas PE 38:2 (*m/z* 770.571) and lysoPI (*m/z* 619.289) were selected as indicative of asymptomatic negative samples (**Figure 4A**). Other species such as PE 50:9 (*m/z* 976.619) and lysoPC 18:1 (*m/z* 556.317) were selected as important for classification and weighted toward symptomatic positive COVID-19 disease for **Classifier 1** and **2** (**Figure 4A and B**). LysoPS 18:1 (*m/z* 522.284) and DG 40:6 (*m/z* 808.504) were selected by Lasso as predictive of COVID-19 positive infection for **Classifier 2** (**Figure 4B**). **Table S3** provides the Lasso features selected for all statistical classifiers, the corresponding identifications, and mass errors.

**Figure 4.**
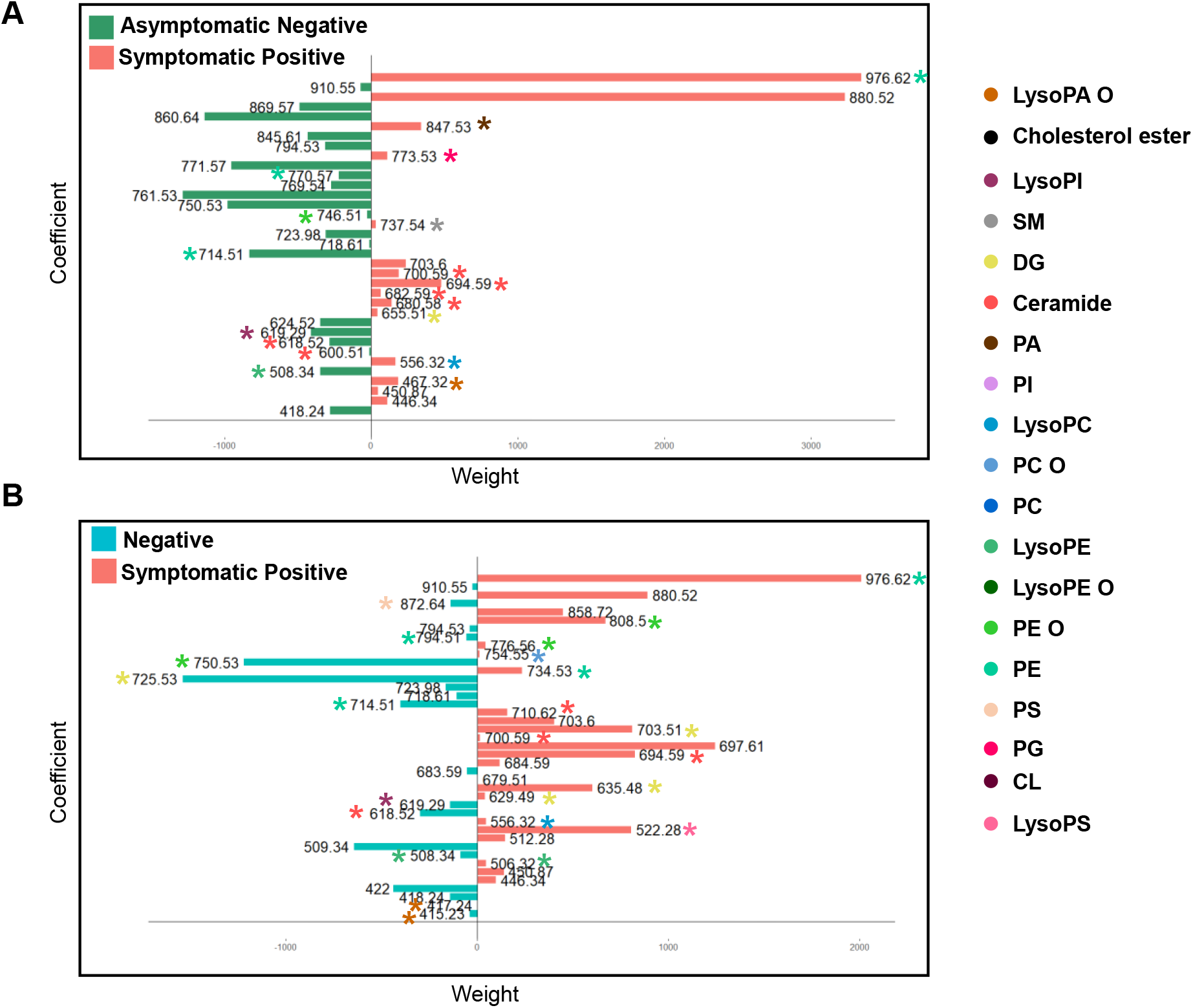
Lasso classification features. Features (*m/z*) selected as indicative of negative infection (negatively weighted values) and positive SARS-CoV-2 infection (positively weighted values) for **A)** Classifier 1 and **B)** Classifier 2. Tentaively identified features are color coded with asterisks corresponding to the identified lipid class.

The results from the descriptive statistical analysis performed to compare the clinical characteristics among the two symptomatic groups, symptomatic COVID-19 PCR positive and COVID-19 PCR negative subjects, are shown in **Table 1**. No association was found among the occurrence of symptoms and comorbidities with the PCR result. These results indicate that there is no clinical difference detected by the symptomatic PCR-positive and symptomatic PCR-negative groups within the patients in our study.

## Discussion

With the slow rollout of the COVID-19 vaccines, a recurring global surge in cases, and the discovery of variants with increased rates of transmission, the availability of alternative technologies that offer rapid analysis and screening for COVID-19 is highly desirable to meet unceasing testing demands. We describe herein the development of a robust MasSpec Pen-ESI MS system for rapid swab analysis and applied the technology to nasopharyngeal swabs in order to evaluate its usefulness for COVID-19 screening.

Modifications were made to the MasSpec Pen design and system to improve the performance and sensitivity for the analysis of swabs. Larger sampling area capabilities were attained to ensure that the three-dimensional clinical swabs with a sparse and heterogeneous distribution of biological materials were sampled in their entirety. The modified PDMS tip includes a hollow middle channel to fit a single swab tip that allowed the full covering and extraction of molecular information from the entire sample during analysis (**Figure 1**), yielding direct, rapid, efficient, and uniform sampling of all the mucous secretion on the swab and largely mitigating bias in the data due to uneven sampling. Additionally, the use of ESI increased the ionization efficiency of extracted lipid molecules and improved the sensitivity for the untargeted molecular analysis of swabs, enabling detection of abundant ions and molecular profiles from the biological sample. Of note, while the total time per analysis of 45 seconds is remarkably fast compared to other available molecular tests, additional system automation approaches are being explored to further expedite device swapping between samples and thus maximize testing throughput. Importantly, similar to the original handheld system, this design of the device for swab analysis maintains a small footprint, retains the ease-of-use and plug-and-play features disposability, and ability to perform rapid molecular analysis, and is compatible with multiple mass spectrometers fitted with ESI interfaces, potentially facilitating implementation in clinical laboratories already equipped with MS instrumentation.

We applied the MasSpec Pen-ESI MS system to analyze 244 nasopharyngeal swabs from COVID-19 positive and negative patients, from which 171 were used to build two statistical classifiers based on the lipid profiles obtained. **Classifier 1** was built to evaluate the performance of our method in discriminating patients diagnosed as positive for COVID-19 infection by PCR from completely asymptomatic patients with a PCR negative result. The classifier was built using 145 samples and yielded a CV prediction accuracy of 83.5%, similar to what achieved in a study using metabolite and lipid information obtained from DESI-MS (86%, n=70) and LD-REIMS (84%, n=70) analysis of heat-inactivated swabs from negative patients with previous SARS-CoV-2 infection and symptomatic positive patients^32^. We evaluated **Classifier 1** on a valdation set of 34 asymptomatic PCR-negative and 14 symptomatic PCR-positive samples, yielding a specificity of 85.3%, or a low FPR of 14.7%, and most notably, a sensitivity of 100%, or low FNR of 0%. These results demonstrate the ability of the MasSpec Pen-ESI technology to detect alterations in the lipid profiles of symptomatic patients with an active SARS-CoV-2 viral infection when compared to healthy individuals and to build classification models based on detected lipid species.

We then used **Classifier 1** to predict on a withheld set samples obtained from 26 symptomatic patients hospitalized with moderate or severe symptoms including fever, cough, difficulty in breathing, but that received a negative PCR result for COVID-19 (**Table S1**). Nine of the symptomatic negative samples had a prediction result in agreement with PCR, while 17 symptomatic negative samples were classified as positive, in disagreement with PCR. Interestingly, 12 of the 17 symptomatic negative samples predicted as positive were obtained from patients showing GGO and consolidations in the chest CT scans, and five of the seven samples predicted as negative presented chest CT results indicative of being negative of viral infection. For example, sample 34 was obtained from a 76 year-old male patient that received a PCR negative diagnosis and was classified as positive by our method. This patient was experiencing cough, sore throat, and dyspnea, and had chest CT features suggestive of infection such as GGO, consolidations, and 50% pulmonary commitment. The patient was hospitalized in the intensive care unit for 13 days with the assistance of a mechanical ventilator until succumbing to death. Additional cases with similar clinical symptoms and results are described in the **Supplementary Information**. Note that chest CT has been suggested as a fundamental tool for early diagnosis and monitoring of COVID-19 as it enables detection of lung alterations in symptomatic patients that are suggestive of viral infection^40-43^. In a recent study, a 90% sensitivity was reported for COVID-19 diagnosis based on GGO combined with other CT features^44^. Yet, chest CT is less specific than PCR and unable to distinguish between an active or previous SARS-CoV-2 infection or a different viral infection causing severe respiratory symptoms. It is also important to note that although PCR is the gold standard for COVID-19 detection and is highly accurate, several studies have reported a sensitivity of 80-90% (or FNR of 10-20%) for COVID-19 diagnosis using nasopharyngeal swabs^45-47^. Thus, while the results obtained on the withheld set of samples indicate that model built using symptomatic PCR positive and asymptomatic PCR negative samples and the selected predictive lipid species are more strongly associated with infection status when used to predict on independent data from symptomatic PCR negative patients, it is also possible that a proportion of the symptomatic PCR negative samples obtained from hospitalized patients may have innacurate PCR results.

In order to more broadly evaluate the ability of our method to identify individuals negative for COVID-19 disease including symptomatic patients, we then built **Classifier 2** using a training set of symptomatic positive samples (n=44) and a negative class (n=127) comprised of data from both symptomatic and asymptomatic PCR negative samples. As a limited number of symptomatic negative samples were used in **Classifier 2**, only a training set of samples was used to assess the performance of the model for COVID-19 screening. Using CV, **Classifier 2** yielded a 78.4% overall agreement with PCR results, 81.8% sensitivity (FNR of 18.2%), and 77.2% specificity (FPR of 22.8%), similar to what achieved for the training set of **Classifier 1** despite the incorporation of symptomatic negative samples into the negative class. Of the 65 samples classified as positive by our method, 36 were from symptomatic positive samples, yielding a PPV value of 55.4%, meaning that for the prevalence of the disease in the cohort of patients evaluated (25.7%), 55.4% of patients with a positive prediction result by our method were also diagnosed as positive for COVID-19 by PCR. However, for the same disease prevalence, a high NPV of 92.5% was achieved, meaning that 92.5% of patients with a negative prediction by our method also received a negative PCR result for COVID-19. Thus, the high NPV value of 92.5% and the FNR of 18.2% achieved provides evidence that **Classifier 2** can potentially identify individuals negative for SARS-CoV-2 infection and predict patients with SARS-CoV-2 infection as positive for the disease, respectively, both of which are paramount to halting the spread of the COVID-19 disease. In comparison, antigen tests aiming to rapid screen for COVID-19 were reported to present mean sensitivity of 64.9% and 51.5% for detection in symptomatic and asymptomatic people, respectively^48^. Here we show that the rapid lipid profiling of nasal swabs can offer improved sensitivity and be used to explore the biochemical information from the host-pathogen interaction. While these results are encouraging, a larger cohort of samples is needed to validate the results by **Classifiers 1** and **2** and further refine and improve the performance and robustness of the model for distinguishing symptomatic COVID-19 positive disease from symptomatic patients with other viral respiratory infections, such as the common cold and influenza.

Our statistical models were based on various classes of phospholipid species previously reported to play key roles in coronavirus virion production and replication^21,22,34^ and that are major components of host cellular membranes. The tentatively identified lipids included glycerophospholipids, such as ceramides, lysolipids, and PE. Among those, several lysoPC species including lysoPC 18:1 were selected as predictive of symptomatic COVID-19 positive disease. Interestingly, in a study by Yan *et al*, lysoPC species were detected at higher abundances in cells infected with the human coronavirus HCoV-229E, when compared to healthy cells, which substantiates our findings^22^. Yet, a recent study by Delafiori *et al* reported decreased abundances of lysoPC species in the serum of COVID-19 positive patients^33^. Across both classifiers, PE 50:9 was selected as indicative of COVID-19 with the highest weight toward the disease, while other PE species such as PE 34:2 was selected as indicative of being negative for COVID-19. Increased abundance of PE species were also recently reported by Ford *et al* in nasal swabs from COVID-19 positive patients^32^. Importantly, since our method does not enable deconvolution between the lipid signal arising from the virion or from host cells, we speculate that the species observed as indicative of COVID-19 are a major component of the virion cellular membrane and/or have increased abundances in host infected cells to enable replication of the virus. Thus, although lipid species represent promising detection targets for COVID-19, additional research is needed to elucidate the role of these species in COVID-19 disease and host response to the infection.

This study has a few limitations. Concerning clinical samples, the swabs for MS and PCR analysis were collected separately for hospitalized patients in our study, which could potentially lead to discrepancies in their diagnoses, especially considering the reported FNR of PCR analysis ^45-47^. Viral load information was also unavailable for the patients which prevented evaluation of a potential relationship between viral burden, molecular information, and diagnostic performance achieved. Heat-inactivation was also used in our study for all the clinical swabs, and thus, biosafety considerations in swab collection, storage, and inactivation steps are needed in future studies to facilitate sample collection and transport. Lastly, although our study was performed using a restricted population of individuals from Brazil, the promising results obtained warrants further investigation, and we expect that a larger cohort of patient samples including asymptomatic PCR positive patients and patients with other viral infections causing similar symptoms to COVID-19 will allow further refinement and validation of the classifiers for COVID-19 disease prediction using lipid information.

In conclusion, the integration of a redesigned version of the disposable MasSpec Pen device provides a rapid MS-based screening method for COVID-19 disease directly from nasopharyngeal swabs. Modifications to the sampling unit and coupling to ESI enabled more effective and reproducible extraction and ionization of lipids from COVID-19 clinical nasal swabs using common solvent systems while maintaining the disposability and user-friendly features of the MasSpec Pen device. As the MasSpec Pen-ESI system has a small footprint and is compatible with various mass spectrometers, this system could be potentially implemented in clinical laboratories and testing facilities that are currently suited with MS instrumentation. The speed of analysis (∼45 s/swab) combined with a relatively lower FNR compared to other FDA approved screening methods and high NPV value achieved substantiate the potential of the technology as a rapid screening tool to identify individuals negative for COVID-19 infection prior to or when PCR is not readily available. While further refinement and testing of the methodology and statistical classifiers with larger sample cohorts will be pursued to improve analytical and diagnostic performance, the present results point to the MasSpec Pen-ESI MS system as a valuable approach for rapid screening of clinical swabs on a seconds-to-minutes time scale.

## Supporting information

Supportin Information

## Data Availability

Raw data is available upon reasonable request

## Acknowledgments

We are grateful to Genio Technologies, Inc., to the Coordination for the Improvement of Higher Education Personnel (CAPES # 88887.504805/2020-00), São Paulo Research Foundation (grant 2019/04314-6), and MackPesquisa (grant 191003) for funding this study. We thank UNIFAG, Santa Casa, and Bragantino Hospitals and their clinical staff for facilitating and contributing to sample collection. We are thankful to every volunteer who accepted to participate in this study. We are also grateful to Tim Hooper for his assistance with the lab-built ESI source.

## Conflict of Interest

K.Y.G., M.F.K., S.C.P., J.Q.L., J.Z., and L.S.E. are inventors in US Patent No. 10,643,832 owned by Board of Regents of the University of Texas System and/or in a provisional patent application related to the MasSpec Pen Technology licensed to MS Pen Technologies, Inc. J.Z. and L.S.E. are shareholders in MS Pen Technologies, Inc. L.S.E. serves as chief scientific officer for MS Pen Technologies, Inc. All other authors declare no competing interests.

## Author contribution

L.S.E., M.N.E., and A. M. P. conceived and designed the research; K.Y.G., M.F.K., J.Z., and L.S.E. designed the MasSpec Pen system; K.Y.G., M.F.K., S.C.P., M.S., J.Z., R.J.D., A.B., and S.B., performed mass spectrometry experiments; A.R.S., J.R.R., P.H.G.S., A.V.M., D.C., and P.H.D.G. collected clinical swabs and clinical data; J.Q.L., J.M.G., K.Y.G, M.F.K., and R.T. performed statistical analysis; L.D.O.N. and M.A.A. performed and supervised clinical research; L.S.E., M.N.E., T.C.C., and A. M. P. provided funding and supervision for the study; K.Y.G., A.M.P, and L.S.E. wrote the manuscript; and all authors revised, read, and approved the final manuscript.

## Data and materials availability

The data used in this study have been reported in Dataverse, https://doi.org/10.7910/DVN/6URYEH

## Notes

### Author Declarations

Approval from Institutional Review Board (IRB) was received for the study (protocol number 31573020.9.0000.5514, Universidade Sao Francisco, approved from May 29, 2020).

