## Supplementary material for "Rapid Screening of COVID-19 Disease Directly from Clinical Nasopharyngeal Swabs using the MasSpec Pen Technology": Supportin Information

**Table of Contents**

**Supplementary Methods**

**Supplementary Figure 1.** Comparison of mass spectra obtained using ESI or solvent-assisted inlet ionization as the ionization mechanism.

**Supplementary Figure 2.** Mass spectra obtained from the MasSpec Pen-ESI analysis of a nylon flock swab containing a lipid standard.

**Supplementary Figure 3.** Comparison of mass spectra acquired from the analysis of swabs with and without heat inactivation.

**Supplementary Table 1.** Demographic and clinical information for symptomatic negative patients in the withheld set for Classifier 1 and training set of Classifier 2.

**Supplementary Table 2.** Overall classification results and agreement with clinical PCR diagnosis for Classifier 1 and 2.

**Supplementary Table 3.** Identification, observed *m/z*, and mass error for ions selected by Lasso as indicative of negative and positive COVID-19 infection.

### Supplementary Methods

#### Engineering the lab-built ESI housing and sprayer

Aluminum blocks and rubber gaskets were machined onto the source to create a vacuum seal when interfaced to the mass spectrometer, and a metal adapter was machined into the bottom of the aluminum block to connect a vacuum tube attached to an external rough pump (Edwards Vacuum). Additionally, a ceramic block was engineered and added to hold the lab-built ESI sprayer. The lab-built sprayer consists of a metal capillary (7 in, 0.62 in OD, 0.16 in ID, New England Small Tube Corporation) concentric with a ceramic tube (10 in, 0.125 in OD, 0.063 in ID, Omega Engineering, Inc.) through which $N_2$ gas flows acting as a sheath gas, a plastic union tee (0.25 in, Legris) to which a gas tube is attached, and two neoprene rubber stoppers each machined with a hole to fit the ceramic tube or metal capillary. A 1.5 m polytetrafluoroethylene (PTFE) tubing and silicone were used to connect the PDMS swab sampling port to the metal capillary of the lab-built sprayer.

#### Handling and storage of clinical nasal swabs

HydraFlock polyester swabs sterilized by gamma irradiation (Puritan Medical Products) were shipped to Brazil. A permit to import samples from Brazil to UT Austin was received from the Brazilian Health Regulatory Agency (ANVISA) and the Center for Disease Control and Prevention (CDC). Prior to shipment, all swabs were heat-inactivated in Brazil at 65°C for 30 min and kept at -20°C until the shipment in dry ice. Experiments were performed at the University of Texas at Austin under Biosafety Level 2 conditions. Swabs were handled by the principal investigator in the biosafety cabinet following biosafety protocols and requirements from the institutional biosafety committee.

#### Optimization of MasSpec Pen-ESI MS system for analysis of swabs

Various swab tip materials were tested using the MasSpec Pen-ESI MS system, including nylon (Copan Diagnostics) and polyester flock (Puritan Medical Products) swabs. Each swab was dipped in a 13 µM standard solution of CL 72:4 and analyzed using the MasSpec Pen-ESI system in the negative ion mode, with the mass spectrum of polyester flock swabs containing minimal interfering ions in the lipid range compared to nylon flock swabs. The interfering ions presented polymer-like distribution and were identified as repeating units of ethylene oxide. Additionally, solvents including methanol, isopropylalcohol, ethanol:ethylacetate (1:1, v/v), $CHCl_3$:MeOH (1:1, v/v), and $CHCl_3$:MeOH (2:1, v/v) were tested for compatibility with the MasSpec Pen-ESI MS system.

To evaluate the robustness of the MasSpec Pen-ESI MS system for the detection of lipids from gamma-irradiated polyester flock swabs, swabs dipped in 100 µL of an equimolar (10 µM) lipid standard mixture of PE 36:2 (*m/z* 742.540) and PG 36:2 (*m/z* 773.539) were analyzed using $CHCl_3$:MeOH (1:1, v/v) as the solvent, yielding a relative standard deviation of 6.4% (n=10).

To evaluate the effect of heat inactivation of the biological material on the swabs, swabs dipped in a mixture of PG and PE lipid standards were heat-treated for 30 min at 65°C (n=5) to simulate heat-inactivation, while control swabs (n=5) dipped in the same lipid mixture standard were not heat-treated, followed by analysis using the MasSpec Pen-ESI system

**MasSpec Pen-ESI MS analysis of clinical nasal swabs**

Extraction solvent, solvent volume, and swab type were optimized using the ion trap analyzer of an LTQ Orbitrap XL, while the heat inactivation experiments, reproducibility experiments, and analysis of clinical swabs were performed using the orbitrap mass analyzer of a Q Exactive HF mass spectrometer. MasSpec Pen-ESI MS analysis of swabs was performed in the negative ion mode at a mass range of *m/z* 100-1500, resolving power of 120,000, and an inlet temperature of 300°C. A voltage of 3.3 kV was applied to the ESI sprayer, and the nitrogen sheath gas pressure was set to 20 psi. The pressure inside the ESI source was approximately 1.4-1.6 mbar for all analyses and experiments. Selected molecular ions were identified by tandem MS and high mass accuracy measurements acquired during MasSpec Pen-ESI MS analyses of swabs on the Q Exactive HF. For the analysis of swabs, chloroform:methanol ($CHCl_3$:MeOH, 1:1, v/v) was used as the extraction solvent. A new sampling device and tubing and a clean sprayer were used for each analysis to avoid cross-contamination between each analysis.

**Statistical analysis**

The reproducibility of the MasSpec Pen-ESI system for swab analysis was quantified by calculating the relative standard deviation using the ratio of the intensities of lipid standards, *m/z* 773.534 (PG 36:2)/*m/z* 742.539 (PE 36:2) (n=10). To evaluate the effect of heat inactivation on the stability of lipids, a t-test was performed on the intensities of two lipid standards, *m/z* 742.639 and *m/z* 773.533, extracted from swabs without heat inactivation (n=5) and swabs heat-inactivated for 30 min at 65°C (n=5) using the MasSpec Pen-ESI system. Lastly, descriptive statistics were also computed for clinical and demographic variables. Categorical variables were expressed as count (percentage), whereas continuous variables were expressed as median (25–75$^{th}$ percentiles). The chi-square test was used to compare categorical variables proportions, and the Mann–Whitney U-test was used to evaluate continuous data.

**Supplementary Discussion**

**Case Studies**

Sample 108 was obtained from a patient with preexisting conditions, including obesity and hypertension. This person received a PCR negative diagnosis and was classified as positive by our method. The symptoms included chest CT results suggestive of infection with GGO, consolidations, crazy-paving appearance, and pulmonary commitment (50%), as well as fever and low $O_2$ saturation levels (<95%). The patient was discharged from the hospital after nine days. Sample 242 was collected from a patitent who received a PCR negative diagnosis and was also classified by our method as negative for viral infection. The patient was hospitalized for three days with symptoms including coughing, sore throat, and dyspnea, whereas her chest CT results were not indicative of an infection.

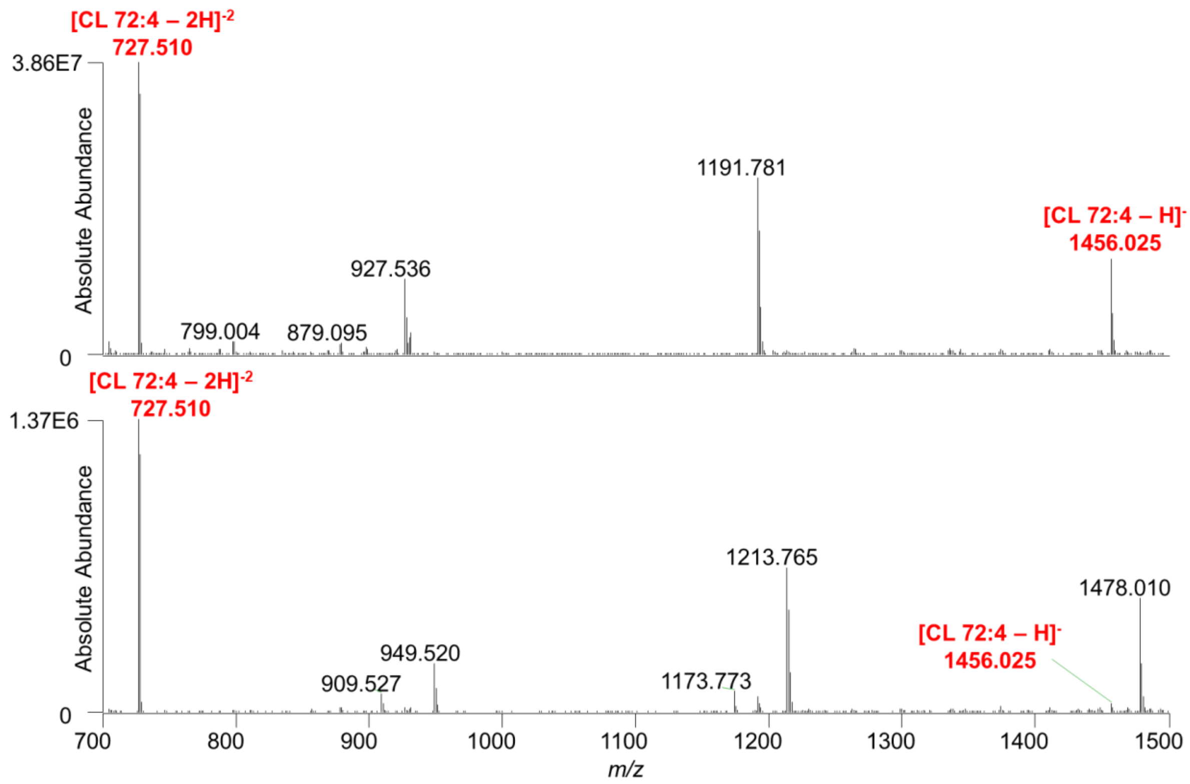


**Supplementary Figure 1.** Comparison of mass spectra obtained using the MasSpec Pen with ESI or solvent assisted inlet ionization to analyze a 20 ppm CL 72:4 lipid standard. The top mass spectrum shows the profile obtained using ESI as the ionization mechanism and the bottom spectrum shows the data collected using a solvent-assisted inlet ionization. The singly and doubly charged CL species are labeled in red.

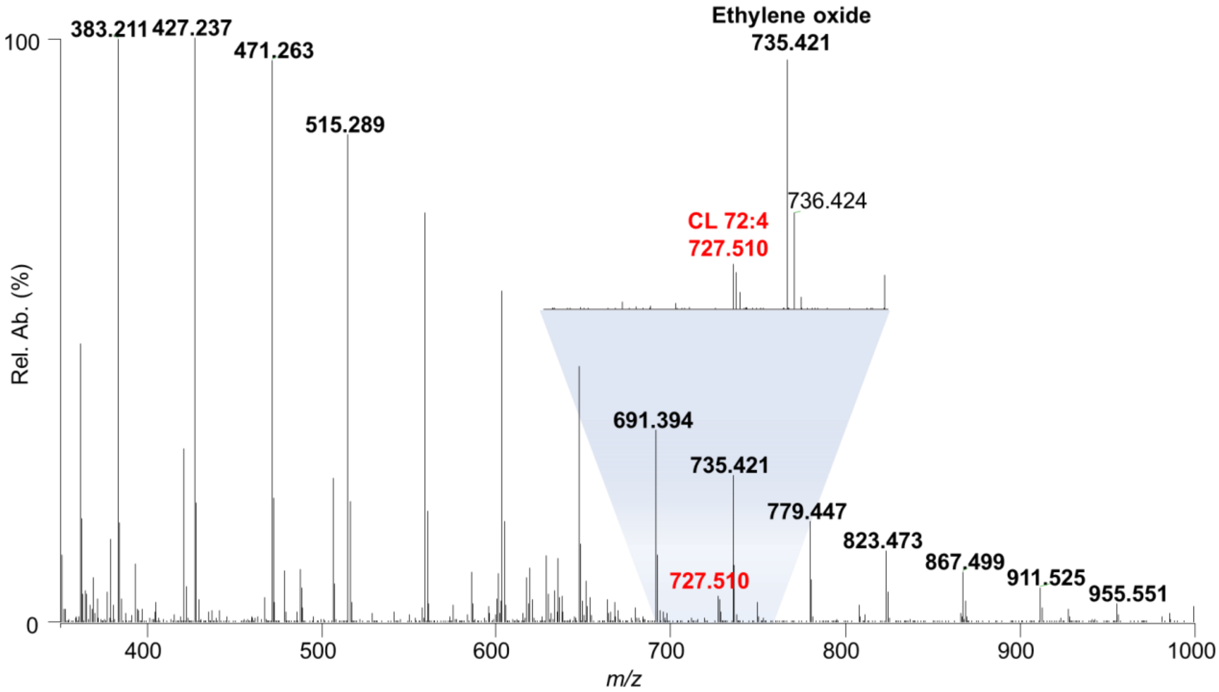


**Supplementary Figure 2.** Representative mass spectrum from the MasSpec Pen-ESI MS analysis of a nylon flock swab dipped in 20 ppm CL 72:4 (*m/z* 727.510) lipid standard. The doubly charged CL species are labeled in red while the repeating units of ethylene oxide are labeled in black.

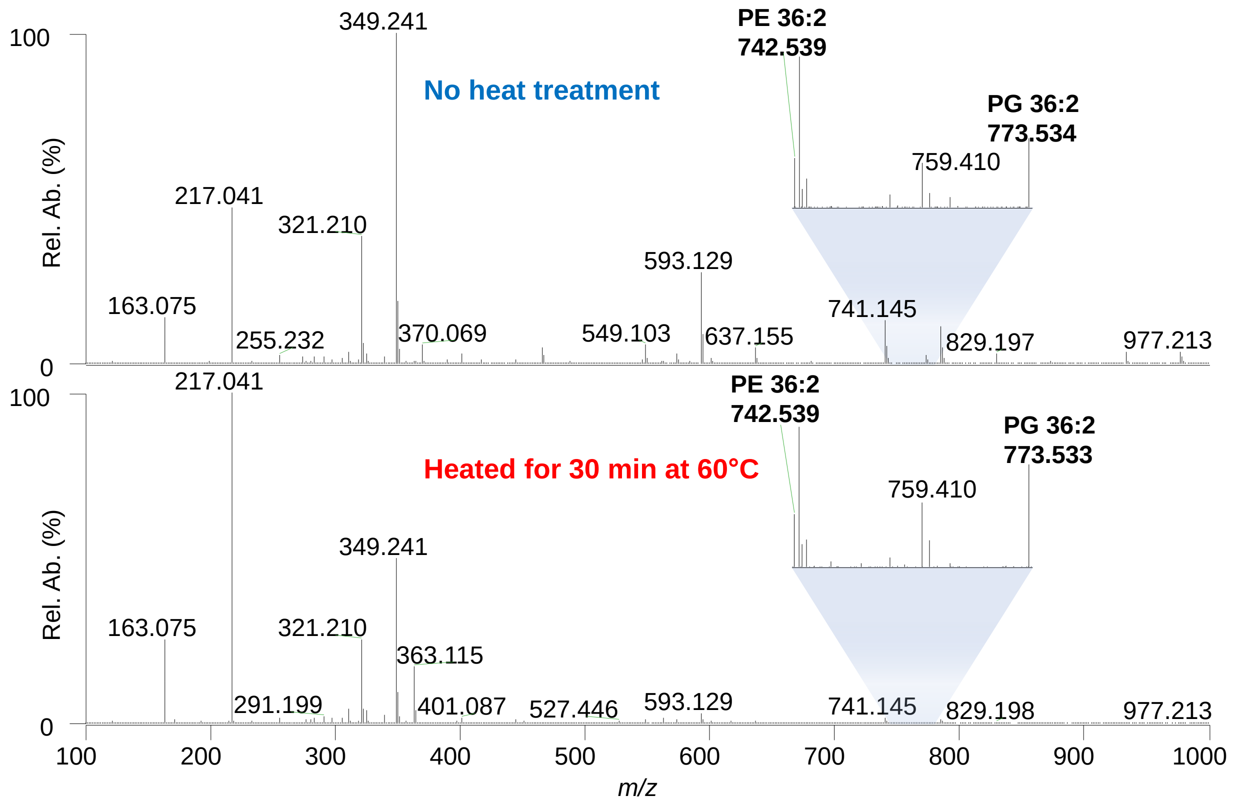


**Supplementary Figure 3** Comparison of mass spectra of swabs containing a 10 μM PG 36:2 (*m/z* 773.533) and PE 36:2 (*m/z* 742.539) lipid standard mixture with and without heat inactivation. The mass spectrum on top was obtained from the MasSpec Pen-ESI MS analysis of a swab without heat inactivation while the bottom shows the profile for a swab that had been heat-inactivated at 60°C for 30 min. Both insets are a zoom of *m/z* 720-800 range to show the detection of the lipid standards.

**Supplementary Table 1.** Demographic and detailed clinical information, including clinical diagnosis, hospitalization, chest CT, and symptom information, for the 26 symptomatic negative patients in the withheld set of samples used in Classifier 1 and in the training set for Classifier 2.

| Original Class | ID# | RT-PCR (Observed) | Prediction (classifier 1) | Hospitalization Days | ICU days | Mechanical Ventilation Days. | Outcome |
|---|---|---|---|---|---|---|---|
| Symptomatic Negative | 30 | NEG | POS | 2 | 0 | 0 | Discharged |
| Symptomatic Negative | 34 | NEG | POS | 15 | 13 | 11 | Death |
| Symptomatic Negative | 36 | NEG | POS | 3 | 0 | 0 | Discharged |
| Symptomatic Negative | 45 | NEG | POS | 13 | 0 | 0 | Discharged |
| Symptomatic Negative | 68 | NEG | POS | 3 | 0 | 0 | Discharged |
| Symptomatic Negative | 71 | NEG | POS | 4 | 0 | 0 | Discharged |
| Symptomatic Negative | 92 | NEG | POS | 29 | 19 | 19 | Death |
| Symptomatic Negative | 93 | NEG | POS | 7 | 0 | 0 | Discharged |
| Symptomatic Negative | 96 | NEG | POS | 5 | 0 | 0 | Discharged |
| Symptomatic Negative | 97 | NEG | POS | 5 | 0 | 0 | Discharged |
| Symptomatic Negative | 101 | NEG | POS | 1 | 0 | 0 | Discharged |
| Symptomatic Negative | 108 | NEG | POS | 9 | 0 | 0 | Discharged |
| Symptomatic Negative | 111 | NEG | POS | 2 | 0 | 0 | Discharged |
| Symptomatic Negative | 116 | NEG | NEG | 8 | 0 | 0 | Discharged |
| Symptomatic Negative | 149 | NEG | POS | 7 | 0 | 0 | Discharged |
| Symptomatic Negative | 151 | NEG | POS | 7 | 0 | 0 | Discharged |
| Symptomatic Negative | 152 | NEG | POS | 4 | 0 | 0 | Death |
| Symptomatic Negative | 156 | NEG | NEG | 3 | 0 | 0 | Discharged |
| Symptomatic Negative | 193 | NEG | POS | 6 | 0 | 0 | Discharged |
| Symptomatic Negative | 240 | NEG | POS | 10 | 0 | 0 | Discharged |
| Symptomatic Negative | 242 | NEG | NEG | 3 | 0 | 0 | Discharged |
| Symptomatic Negative | 263 | NEG | POS | 6 | 0 | 0 | Discharged |
| Symptomatic Negative | 264 | NEG | POS | 2 | 0 | 0 | Discharged |
| Symptomatic Negative | 265 | NEG | NEG | 3 | 0 | 0 | Discharged |
| Symptomatic Negative | 266 | NEG | POS | 2 | 0 | 0 | Discharged |
| Symptomatic Negative | 267 | NEG | NEG | 64 | 0 | 0 | Discharged |

| Original Class | ID# | Chest CT Suggestive of viral infection | Chest CT Findings | | | | | |
|---|---|---|---|---|---|---|---|---|
| | | | Ground-Glass Opacity | Consolidations | Crazy Paving Appearance | Reticular Pattern | resence of Pulmonary Commitme | % of Pulmonary Commitment |
| Symptomatic Negative | 30 | YES | YES | NO | NO | NO | YES | 25-50 |
| Symptomatic Negative | 34 | YES | YES | YES | NO | NO | YES | 50 |
| Symptomatic Negative | 36 | YES | YES | NO | YES | YES | YES | 20-50 |
| Symptomatic Negative | 45 | YES | YES | YES | YES | YES | YES | 50 |
| Symptomatic Negative | 68 | NO | NO | NO | NO | NO | NO | 0 |
| Symptomatic Negative | 71 | YES | YES | YES | NO | NO | YES | 50 |
| Symptomatic Negative | 92 | YES | YES | NO | NO | NO | YES | 30-50 |
| Symptomatic Negative | 93 | YES | YES | YES | YES | NO | YES | 60 |
| Symptomatic Negative | 96 | YES | YES | YES | NO | YES | YES | 20-50 |
| Symptomatic Negative | 97 | YES | YES | YES | YES | YES | YES | 30 |
| Symptomatic Negative | 101 | YES | YES | YES | NO | NO | NO | 0 |
| Symptomatic Negative | 108 | YES | YES | YES | YES | YES | YES | 50 |
| Symptomatic Negative | 111 | YES | YES | YES | NO | NO | YES | 50-80 |
| Symptomatic Negative | 116 | NO | NO | NO | NO | NO | NO | 0 |
| Symptomatic Negative | 149 | YES | YES | YES | NO | NO | YES | 50-70 |
| Symptomatic Negative | 151 | YES | YES | YES | YES | NO | YES | 50-80 |
| Symptomatic Negative | 152 | YES | YES | YES | YES | NO | YES | 50 |
| Symptomatic Negative | 156 | NO | NO | NO | NO | NO | NO | - |
| Symptomatic Negative | 193 | YES | YES | YES | YES | NO | YES | 50-80 |
| Symptomatic Negative | 240 | NO | NO | NO | NO | NO | NO | 0 |
| Symptomatic Negative | 242 | NO | NO | NO | NO | NO | NO | 0 |
| Symptomatic Negative | 263 | NO | NO | YES | NO | NO | NO | 0 |
| Symptomatic Negative | 264 | YES | YES | NO | NO | NO | NO | 0 |
| Symptomatic Negative | 265 | NO | NO | NO | NO | NO | NO | 0 |
| Symptomatic Negative | 266 | NO | NO | NO | NO | NO | NO | 0 |
| Symptomatic Negative | 267 | YES | YES | NO | YES | YES | YES | 20-50 |

| Original Class | ID# | Reported Symptons | | | | | | | | | Other information | | |
|---|---|---|---|---|---|---|---|---|---|---|---|---|---|
| | | Fever | Cough | Myalgia | Sore.Throat | Headache | Coryza | Dyspnea | Loss of smell/taste | O2 Sauration < 95% | COPD presence | Smoker or Ex-smoker | Asthma |
| Symptomatic Negative | 30 | YES | YES | NO | NO | NO | NO | YES | YES | YES | NO | NO | NO |
| Symptomatic Negative | 34 | NO | YES | NO | YES | NO | NO | YES | NO | NO | NO | NO | NO |
| Symptomatic Negative | 36 | YES | NO | NO | NO | NO | NO | YES | NO | YES | NO | NO | NO |
| Symptomatic Negative | 45 | NO | NO | YES | NO | NO | NO | YES | NO | NO | NO | NO | NO |
| Symptomatic Negative | 68 | YES | NO | NO | NO | NO | NO | YES | YES | YES | NO | NO | NO |
| Symptomatic Negative | 71 | YES | NO | NO | YES | NO | NO | NO | YES | YES | NO | NO | NO |
| Symptomatic Negative | 92 | NO | YES | NO | NO | NO | NO | YES | NO | NO | NO | NO | NO |
| Symptomatic Negative | 93 | NO | YES | NO | NO | NO | NO | YES | YES | NO | NO | NO | NO |
| Symptomatic Negative | 96 | NO | YES | NO | NO | NO | NO | YES | NO | NO | YES | YES | NO |
| Symptomatic Negative | 97 | YES | NO | NO | YES | NO | YES | NO | NO | NO | NO | NO | NO |
| Symptomatic Negative | 101 | YES | NO | NO | NO | NO | NO | NO | NO | YES | NO | NO | YES |
| Symptomatic Negative | 108 | NO | YES | NO | NO | NO | NO | NO | NO | YES | NO | NO | NO |
| Symptomatic Negative | 111 | NO | YES | NO | NO | NO | NO | NO | YES | YES | NO | NO | NO |
| Symptomatic Negative | 116 | NO | NO | NO | NO | NO | NO | YES | YES | NO | NO | NO | NO |
| Symptomatic Negative | 149 | YES | NO | NO | YES | NO | NO | NO | NO | NO | NO | NO | NO |
| Symptomatic Negative | 151 | NO | YES | NO | YES | YES | NO | YES | YES | YES | NO | NO | NO |
| Symptomatic Negative | 152 | YES | NO | NO | NO | NO | NO | YES | YES | NO | NO | YES | NO |
| Symptomatic Negative | 156 | NO | YES | NO | YES | NO | NO | YES | NO | NO | NO | NO | YES |
| Symptomatic Negative | 193 | YES | NO | YES | NO | NO | NO | NO | YES | YES | NO | NO | NO |
| Symptomatic Negative | 240 | YES | YES | NO | NO | NO | YES | YES | NO | NO | NO | YES | NO |
| Symptomatic Negative | 242 | NO | YES | NO | YES | NO | YES | NO | NO | NO | NO | NO | NO |
| Symptomatic Negative | 263 | YES | YES | NO | NO | NO | NO | YES | NO | NO | NO | NO | NO |
| Symptomatic Negative | 264 | YES | YES | NO | YES | NO | NO | YES | NO | NO | NO | NO | NO |
| Symptomatic Negative | 265 | YES | YES | NO | NO | NO | NO | YES | NO | NO | NO | NO | NO |
| Symptomatic Negative | 266 | YES | YES | NO | NO | NO | NO | YES | NO | NO | NO | NO | NO |
| Symptomatic Negative | 267 | NO | YES | NO | NO | NO | NO | YES | NO | NO | NO | NO | NO |

**Supplementary Table 2.** Confusion matrices of the Lasso results for Classifiers 1 and 2.

| Classification Model | Sample Set | PCR Diagnosis | Model Prediction and Performance | | | | | | | | |
|---|---|---|---|---|---|---|---|---|---|---|---|
| | | | **Negative** | **Positive** | **Sensitivity (%)** | **Specificity (%)** | **Accuracy (%)** | **NPV (%)** | **PPV (%)** | **FNR** | **FPR** |
| **Classifier 1: Asymptomatic Negative vs Symptomatic Positive** | Training | Negative | 58 | 9 | 76.7 | 86.6 | 83.5 | 89.2 | 71.9 | 23.3 | 13.4 |
| | | Positive | 7 | 23 | | | | | | | |
| | Validation | Negative | 29 | 5 | 100.0 | 85.3 | 89.6 | 100.0 | 73.7 | 0.0 | 14.7 |
| | | Positive | 0 | 14 | | | | | | | |
| | Withheld Data | Negative | 17 | 9 | - | 65.4 | - | - | - | | |
| **Classifier 2: Asymptomatic + Symptomatic Negative vs Symptomatic Positive** | Training | Negative | 98 | 29 | 81.8 | 77.2 | 78.4 | 92.5 | 55.4 | 18.2 | 22.8 |
| | | Positive | 8 | 36 | | | | | | | |

**Supplementary Table 3.** Observed *m/z*, mass error, and identification for the features selected by Lasso for Classifier 1 and 2. Identifications are based on high mass accuracy and/or tandem MS measurements.

| Lasso Feature | Observed *m/z* | Theoretical *m/z* | Mass Error | Attribution | Formula |
|---|---|---|---|---|---|
| 415.23 | 415.226 | 415.2255 | 1.204 | LPA O-18:4 | C21H37O6P [M-H]- |
| 417.24 | 417.241 | 417.2412 | -0.479 | LPA O-18:3 | C21H39O6P [M-H]- |
| 418.24 | 418.244 | 418.2445 | -1.195 | Isotope of LPA O-18:3, m/z 417.241 | C21H39O6P [M-H]- |
| 422 | 421.997 | - | - | Unidentified | - |
| 446.34 | 446.336 | - | - | Isotope of m/z 445.333, unidentified | - |
| 450.87 | 450.874 | - | - | Unidentified | - |
| 467.32 | 467.316 | 467.3143 | 3.638 | LPA O-20:0 | |
| 506.32 | 506.324 | 506.3252 | -2.370 | LPE 20:1 | C25H50NO7P [M-H]- |
| 508.34 | 508.341 | 508.3409 | 0.197 | LPE 20:0 | C25H52NO7P [M-H]- |
| 509.34 | 509.344 | 509.3442 | -0.393 | Isotope of LPE 20:0 | C25H52NO7P [M-H]- |
| 512.28 | 512.283 | 512.2864 | - | Unidentified | - |
| 522.28 | 522.284 | 522.2838 | 0.383 | LPS 18:1 | C24H46NO9P [M-H]- |
| 556.32 | 556.318 | 556.3176 | 0.719 | LPC 18:1 | C26H52NO7P |
| 600.51 | 600.514 | 600.5128 | 1.998 | Cer 36:1 | C36H71NO3 [M+Cl]- |
| 618.52 | 618.524 | 618.5234 | 0.970 | Cer 36:0 | C36H73NO4 [M+Cl]- |
| 619.29 | 619.289 | 619.2889 | 0.161 | LPI 20:4 | C29H49O12P [M-H]- |
| 624.52 | 624.522 | - | - | Unidentified | - |
| 629.49 | 629.492 | 629.4917 | 0.477 | DG 34:1 | C37H70O5 [M+Cl]- |
| 635.48 | 635.482 | 635.4812 | 1.259 | DG O-36:5 | C39H68O4 [M+Cl]- |
| 655.51 | 655.508 | 655.5074 | 0.915 | DG 36:2 | C39H72O5 [M+Cl]- |
| 679.51 | 679.507 | 679.5074 | -0.589 | DG O-38:5 | C41H72O5 [M+Cl]- |
| 680.58 | 680.576 | 680.5754 | 0.882 | Cer 42:3 | C42H79NO3 [M+Cl]- |
| 682.59 | 682.591 | 682.5911 | -0.147 | Cer 42:2 | C42H81NO3 [M+Cl]- |
| 683.59 | 683.594 | 683.5944 | -0.585 | Isotope of m/z 682.591, Cer 42:2 | C42H81NO3 [M+Cl]- |
| 684.59 | 684.588 | 684.5881 | -0.146 | Isotope of m/z 682.591, Cer 42:3 | C42H81NO4 [M+Cl]- |
| 694.59 | 694.592 | 694.5911 | 1.296 | Cer 43:3 | C43H81NO3 [M+Cl]- |
| 697.61 | 697.61 | - | - | Unidentified | - |

| | | | | | |
|---|---|---|---|---|---|
| 700.59 | 700.587 | 700.5886 | -2.284 | Cer 44:5 | C44H79NO5 [M+Cl]- |
| 703.51 | 703.507 | 703.5074 | -0.569 | DG 40:6 | C43H72O5 [M+Cl]- |
| 703.6 | 703.603 | - | - | Unidentified | - |
| 710.62 | 710.623 | 710.6224 | 0.844 | Cer 44:2 | C44H85NO3 [M+Cl]- |
| 714.51 | 714.508 | 714.5079 | 0.140 | PE 34:2 | C39H74NO8P [M-H]- |
| 718.61 | 718.613 | - | - | Unidentified | - |
| 723.98 | 723.981 | 723.9805 | 0.691 | Isotope of m/z 723.479, CL 72:8 | C81H142O17P2 [M-2H]2- |
| 725.53 | 725.533 | 725.5362 | -4.411 | DG 42:7 | C45H74O7 [M-H]- |
| 734.53 | 734.534 | 734.5342 | -0.272 | PE 34:0 | C39H78NO9P [M-H]- |
| 737.54 | 737.537 | 737.5370 | 0.000 | SM 34:1 | C39H79N2O6P [M+Cl]- |
| 746.51 | 746.513 | 746.5130 | 0.000 | PE O-38:7 | C43H74NO7P [M-H]- |
| 750.53 | 750.528 | 750.5292 | -1.599 | HexCer 34:1 | C49H77NO9 [M+Cl]- |
| 754.55 | 754.554 | 754.5523 | 2.253 | PC O-32:0 | C40H82NO7P [M+Cl]- |
| 761.53 | 761.534 | 761.532 | 2.626 | Isotope of m/z 760.529, PE O-39:7 | C44H76NO7P [M-H]- |
| 769.54 | 769.535 | 769.5389 | - | Unidentified | - |
| 770.57 | 770.570 | 770.5705 | -0.649 | PE 38:2 | C43H82NO8P [M-H]- |
| 771.57 | 771.573 | 771.574 | -1.296 | Isotope of m/z 770.570, PE 38:2 | C43H82NO8P [M-H]- |
| 773.53 | 773.534 | 773.5338 | 0.259 | PG 36:2 | C42H79O10P [M-H]- |
| 776.56 | 776.559 | 776.5600 | -1.288 | PE O-40:6 | C45H80NO7P [M-H]- |
| 794.51 | 794.508 | 794.5108 | -3.524 | PE 36:2 | C41H78NO9P [M+Cl]- |
| 794.53 | 794.529 | - | - | Unidentified | - |
| 808.5 | 808.504 | 808.5054 | -1.732 | PE O-40:8 | C45H76NO7P [M+Cl]- |
| 845.61 | 845.610 | 845.611 | -1.183 | Isotope of m/z 844.607, PS 40:1 | C46H88NO10P [M-H]- |
| 847.53 | 847.529 | 847.5283 | 0.826 | PA 48:12 | C51H77O8P [M-H]- |
| 858.72 | 858.723 | - | - | Unidentified | - |
| 860.64 | 860.638 | 860.6388 | -0.930 | HexCer 42:2 | C48H91NO9 [M+Cl]- |
| 869.57 | 869.569 | 869.5662 | 3.220 | Isotope of m/z 868.563, PC 40:6 | C48H84NO8P [M+Cl]- |
| 872.64 | 872.638 | 872.6386 | -0.688 | PS 42:1 | C48H92NO10P [M-H]- |
| 880.52 | 880.519 | 880.5195 | -0.568 | HexCer 32:3 | C44H79NO14 [M+Cl]- |
| 910.55 | 914.584 | 914.5845 | -0.547 | Isotope of m/z 913.585, PI 40:4 | C49H87O13P [M-H]- |
| 976.62 | 976.619 | 976.6204 | -1.434 | PE 50:9 | C55H92NO9P [M+Cl]- |
